# A New Extension of State-Space SIR Model to Account for Underreporting- An Application to the COVID-19 transmission in California and Florida

**DOI:** 10.1101/2020.12.20.20248580

**Authors:** Vishal Deo, Gurprit Grover

## Abstract

In the absence of sufficient testing capacity for COVID-19, a substantial number of infecteds are expected to remain undetected. Since the undetected cases are not quarantined, they are expected to transmit the infection at a much higher rate than their quarantined counterparts. That is, under the lack of extensive random testing, the actual prevalence and incidence of the SARS-CoV-2 infection may be entirely different from that being reported. Thus, it is imperative that the information on the percentage of undetected (or unreported) cases be considered while estimating the parameters and forecasting the transmission dynamics of the epidemic.

In this paper, we have developed a new version of the basic susceptible-infected-removed (SIR) compartmental model, called the susceptible-infected (quarantined/ free) -recovered-deceased [SI(Q/F)RD] model, to incorporate the impact of undetected cases on the transmission dynamics of the epidemic. Further, we have presented a Dirichlet-Beta state-space formulation of the SI(Q/F)RD model for the estimation of its parameters using posterior realizations from Gibbs sampling procedure. As a demonstration, the proposed methodology is implemented to forecast the COVID-19 transmission in California and Florida.

**Highlights:**

- Data calibrated for underreporting using excess deaths and case fatality rate.
- A new extension of SIR compartmental model, called SI(Q/F)RD, is introduced.
- A Dirichlet-Beta state-space formulation of the SI(Q/F)RD model is developed.
- Gibbs sampling used to estimate the Bayesian hierarchical state-space model.
- Proposed methodology is applied on the COVID-19 data of California and Florida.

## 1. Introduction

As per the scientific brief of the World Health Organization (WHO) published on its website on 9 July 2020, transmission of SARS-CoV-2 occurs primarily between people through direct, indirect, or close contact with infected people through infected secretions such as saliva and respiratory secretions, or through their respiratory droplets, which are expelled when an infected person coughs, sneezes, talks or sings; refer [1]. That is, in order to break the chains of transmissions of SARS-CoV-2, the objective of the preventive measures should be to minimize the contact of susceptibles with infected people. The first step towards this goal is to identify the infecteds so that they can be kept in quarantine till they are no longer infectious. However, high variability in the level and the nature of symptoms in infecteds, coupled with a significant length of incubation period, poses a difficult challenge to frame a targeted testing strategy which can serve the purpose effectively. Although contract tracing can help in identifying the chains of transmission associated with detected cases, presence of a high proportion of asymptomatic cases, also capable of transmitting infection [2, 3], flags concerns about the reliability of the strategy. Further, in the presence of high proportion of asymptomatic cases, limiting testing to the symptomatic individuals will also fail to serve the objective of detecting and quarantining all infecteds. So, apart from testing symptomatic individuals (mild or severe), and identifying and testing high risk individuals having history of contact with infected people, the situation demands aggressive random testing to isolate even the asymptomatic cases from the population. In the absence of adequate amount of random testing, a significant number of infecteds, especially asymptomatic individuals, may remain undetected (*i.e*., number of cases will remain significantly underreported). Since the undetected cases are not quarantined, they are expected to remain infectious in the population for a relatively much longer as compared to those who are detected and quarantined. Lack of any visible symptom in the undetected cases increases the likelihood of susceptibles spending prolonged period in their proximity. Thus, the undetected cases can exhibit a strikingly higher reproduction rate as compared to that of their quarantined counterparts. To be precise, aggressive random testing is imperative towards fulfilling the objective of breaking the chains of transmissions of SARS-CoV-2 using non-medical containment measures.

Despite repeated appeals and advisories to all countries from the WHO to employ extensive random testing, only few countries have shown intentions to conduct adequate number of COVID-19 tests [4]. Rate of positive tests at a place can give an indication about the adequacy, or inadequacy, of the number of tests being carried out in the region. New Zealand has set an extraordinary example in quickly containing the epidemic by efficiently adhering to the strategy of aggressive testing and isolation of infecteds to break the chains of infection. As on 25 August 2020, New Zealand has reported a total of 1690 confirmed cases of which 1539 have recovered, 129 are active and 22 have died. These are very encouraging figures, especially when compared with those of the countries like USA, Brazil, UK, India, Russia, and many others. Till 19 August 2020, the overall percentage of positive tests in New Zealand is reported to be 0.23% [5]. While in USA, the percentage of positive tests has remained quite high since the beginning, with the overall positive percentage staring at 9%. Also, there is a lot of variation between the percentages of positive tests reported by different states in the United States, which varies between 0.53% (Vermont) to 100% (Washington) as reported on 26 August 2020 by the Johns Hopkins University [6]. As per the recommendation of the WHO, the rates of positivity in testing should remain at 5% or lower for at least 14 days at a stretch before governments decide about relaxing the containment/ lockdown measures. This advisory from the WHO is based on the rationale that very high positive rates may indicate that people with only severe symptoms are getting tested and all the asymptomatic cases or cases with mild symptoms are being left out. That is, a high rate of positivity may imply that the testing capacity of the state/ country is insufficient to gauge the actual size of the outbreak, leading to a significant proportion of cases being unreported. As of now, underreporting of cases in the USA has been confirmed and reported by few published scientific studies. Wu et al. [7] have used a semi-Bayesian probabilistic bias analysis to account for incomplete testing and imperfect diagnostic accuracy. As per their estimate, there were 6,454,951 true cumulative infections compared to 721,245 confirmed cases (reported) as of 18 April 2020 in the United States. For different states, the actual number of infections was reported to be 3 to 20 times higher than the confirmed cases. Lau et al. [8] used a simple intuitive method based on crude case fatality risk and adjusted case fatality risk to evaluate extent of underreporting in various COVID-19 epicentres across the world. Their study was based on early-stage data of COVID-19 and they reported severe underreporting of cases in most of the countries worldwide. For USA, they found the estimate of actual number of infections to be around 53.8 times the reported number of confirmed cases. This extraordinarily high estimate of underreporting can be explained by the fact that their study was based on the data reported till 17 March 2020. In general, underreporting is expected to be high at the initial stage of any epidemic owing to the lack of proper system at place, and lack of knowledge and awareness about the infection. Citing their findings, they suggested that due to limited testing capacities, mortality numbers may serve as a better indicator for COVID-19 case spread in many countries.

Although some studies have been done so far to estimate true burden of the disease, the literature lacks any attempt at forecasting true trajectory of infection by dynamically accounting for underreporting of cases. For example, the two papers cited above [7, 8] have been able assess the extent of underreporting till a fixed past date, but they have not presented any methodology to predict the true number of cases under the dynamically changing rates of underreporting. In this paper, we have developed a comprehensive methodology to estimate the true parameters of an epidemic and forecast its transmission dynamics under serious underreporting of cases. To materialize this objective, we propose a new compartmentalised epidemic model called the susceptible-infected (quarantined/ free) - recovered-deceased [SI(Q/F)RD] model. To induce uncertainties in this mathematical model, we have developed its Dirichlet-Beta state-space (Bayesian hierarchical) formulation. The Bayesian hierarchical formulation is implemented in JAGS through the R package ‘R2Jags’ to obtain posterior estimates of the unknown parameters. To demonstrate the implementation of the methodology, we have considered the cases of two of the worst COVID-19 affected states of USA, California, and Florida, which have very high percentages of positive tests. Since the level of testing, and protocols/ procedure of reporting of number of deaths may vary between different state jurisdictions, the level of underreporting of deaths and cases can also be expected to vary between states. This is the reason that we have performed state-wise analyses, rather than analysing the combined data of USA. It should be noted that, although underreporting of cases can occur because of various other reasons, like poor communication between state and hospitals, conscious data manipulation to conceal failures of administration, anomalies in protocols used for declaring epidemic related deaths, lack of proper digital infrastructure to keep reliable records, to name a few, we have assumed that lack of sufficient testing is the sole (or at least major) reason for underreporting. This assumption holds for a developed country like USA, where other reasons like lack of proper communication or digital infrastructure can be conveniently crossed off.

## 2. Methodology

To realize the objective of our study, we propose the following sequence of steps, which are then implemented on the COVID-19 time-series data of California and Florida.

### 2.1. Calibrating daily number of deaths using weekly excess deaths due to COVID-19

Underreporting of deaths has been estimated using the estimates of excess deaths. Excess deaths due to an epidemic (COVID-19 in our case) can be estimated as the difference between the total number of deaths reported in the period of epidemic (from all causes) and the expected number of baseline deaths due to all other causes in the absence of COVID-19. One popular method to calculate the expected number of baseline deaths in the absence of COVID-19 is to fit a Poisson regression to the time-series (weekly) data of death counts, and then projecting the baseline death counts till the required future point in time. An over-dispersed Poisson generalized linear models with spline terms is used to model trends in counts, accounting for seasonality; refer [9-11]. The model is also adjusted for year-to-year baseline variation and any pre-existing epidemic, like influenza epidemic. In our study we have used weekly estimates of excess deaths published by the Centers for Disease Control and Prevention (CDC) for the two states under consideration for the analyses [9]. The estimated excess death counts in the presence of COVID-19 are taken as the estimates of actual (or true) number of deaths due to the pandemic. The reported daily deaths due to COVID-19 are summed over weeks to find weekly reported deaths. The difference between the weekly excess deaths and weekly reported deaths give us the estimate of the unreported deaths due to the pandemic. For further analysis, the weekly estimate of unreported deaths due to COVID-19 is distributed among each day of the corresponding week as per the proportion of the number of pandemic related deaths reported on a day out of the total number of deaths due to the pandemic reported in that week. If all days of a week have zero reported deaths, the total number of unreported deaths is equally distributed among all seven days. Combining the additional death counts assigned to each day to the already reported death counts for the day gives us the calibrated daily time-series data on actual number of deaths due to COVID-19. The calibrated data is smoothed using the method of LOESS regression before proceeding with further calculations.

### 2.2. Estimation of underreporting of number of infecteds

Actual daily number of infecteds can be estimated using a reliable estimate of case fatality rate (CFR). As the reported data on number of cases is expected to suffer from underreporting, CFR based on the population level data will be misleading. If we assume that the level (or proportion) of underreporting of deaths and infecteds are same, the CFR estimated from the reported data will be a reliable estimate of the true population CFR. However, this is hardly the case, and the proportions of underreporting of deaths and infecteds usually differ considerably. An estimate of CFR based on an individual patient level (follow-up) data is deemed as most reliable [12]. So, a simple rule of thumb to know if the levels of underreporting of deaths and infecteds can be assumed to be same is to compare the delayed CFR obtained from reported data with the one obtained from the individual patient level data. If they vary significantly, we should infer that the levels of underreporting are different for number of deaths and number of infecteds. In such a situation, it is advisable to use the CFR obtained from the individual patient level data as the best estimate for the true CFR of the epidemic. Once a reliable estimate of the CFR is obtained, it can be used to calibrate the data for the number of true infecteds on each day using the estimated counts of true deaths and the average delay between infection and death. That is, if the average duration between infection (or detection/ reporting of infection) and death associated with COVID-19 is known to be, say, *h* days, and suppose that *D*_*t*_ number of people have died of the infection on a particular day *t*, then *I*^*n*^_*t-h+1*_ *=* (*D*_*t*_ */* CFR) number of new cases are expected to be infected *h* days prior to the day of death.

If daily reported data on the number of recovered cases (***R***_***t***_) is available, it can be inflated according to the proportion of underreporting estimated in the number of infecteds as follows.

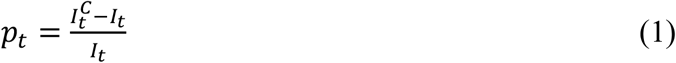

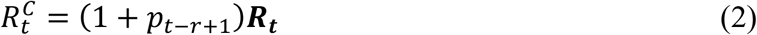

where, *p*_*t*_ is the estimated proportion of underreporting of infecteds at time *t*, 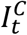 is the calibrated number of infecteds at time *t, I*_*t*_is the reported number of infecteds at time *t*, 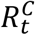 is the calibrated number of recovered cases at time *t*, ***R***_***t***_ is the reported number of recovered cases at time *t* and *r* is the average duration from infection to recovery of patients. If the daily number of recovered cases is not reported, or if it is not reliable, a viable calibration can be done using *r* and (1-CFR). Calculations will be like that used for the calibration of number of infecteds. Further, sum of calibrated counts of total infecteds, total recovered and total deaths till a day will give us the estimate of the cumulative number of confirmed cases till that day.

### 2.3. The proposed SI(Q/F)RD epidemic model

This variation of the popular Susceptible-Infected-Removed (SIR) compartmental model is designed to account for underreporting and its impact on the trajectory of the epidemic. Underreporting is assumed to be the result of some infected cases not getting detected because of the lack of adequate testing capacity. That is, a proportion of the infecteds are detected (*p*) and quarantined (mostly symptomatic cases), while the rest of the infecteds (mostly asymptomatic cases) are undetected and roam freely among the susceptibles. This leads to the belief that the undetected cases are expected to infect the susceptibles at a higher rate (*β*_*2*_), than that of their quarantined counterparts (*β*_*1*_). The proportion of detected cases, *p*, can vary with time if testing capacity is increased or decreased over the period of the epidemic, and can be taken as a function of time *t*, say, *p*_*t*_. The overall structure of this model is presented in Figure 1. Since the quarantined infecteds consist mostly of symptomatic cases, including critical cases, quarantined infecteds can be expected to be at a higher risk of death on an average. Consequently, different death rates can be assumed for quarantined and undetected cases. Different recovery rates can be assumed for quarantined and undetected cases if any scientific evidence exists in its favour. Otherwise, we can assume that both groups of infecteds have equal average recovery rate.

**Figure 1:**
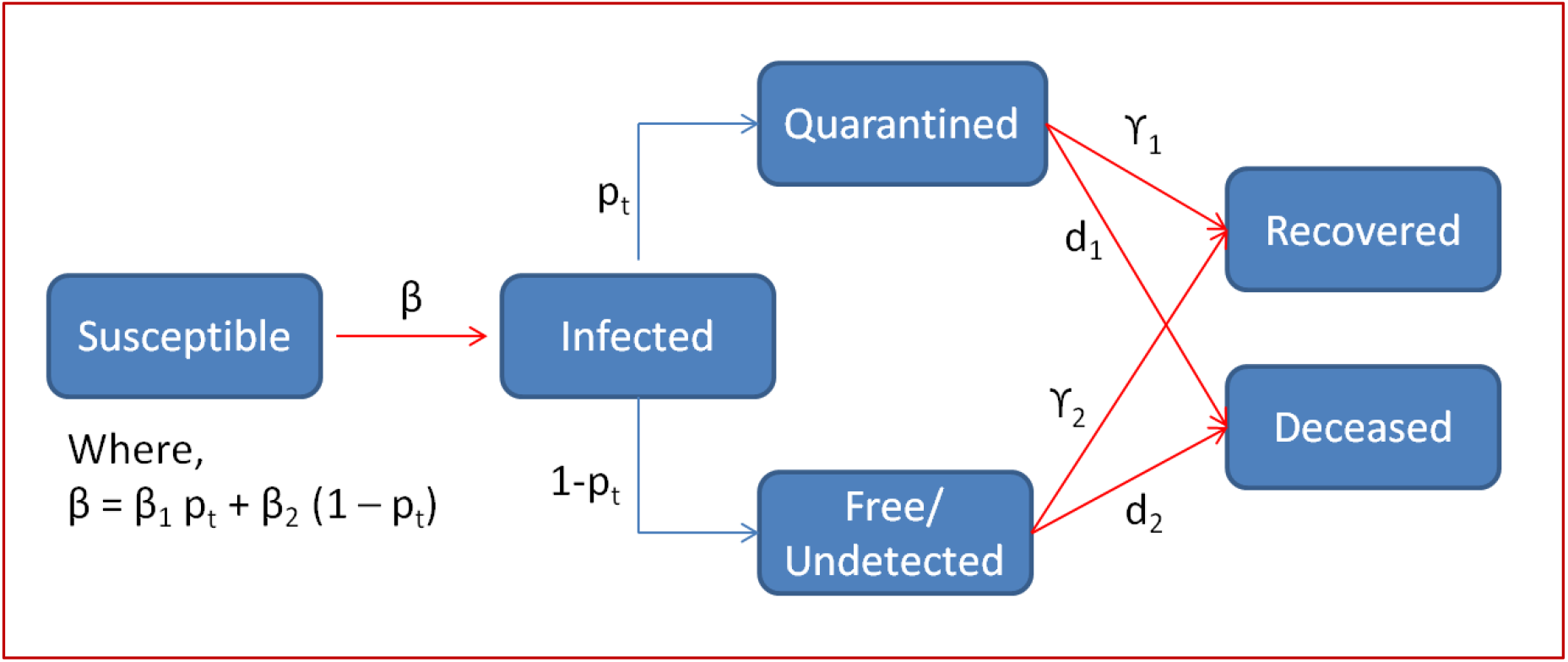
**SI(Q/F)RD model structure-*p*_*t*_ is the proportion of infecteds detected and quarantined, *1-p*_*t*_ is the proportion of infecteds who are undetected and roaming freely among the susceptibles, *β*_*1*_ is the transmission rate associated with quarantined infecteds and *β*_*2*_ is the transmission rate associated with undetected infecteds, *γ*_*1*_ and *d*_*1*_ are rate of recovery and rate of death for quarantined cases and *γ*_*2*_ and *d*_*2*_ are rate of recovery and rate of death for undetected cases.**

The set of differential equations quantifying the transitions defined in Figure 1 can be expressed as follows.

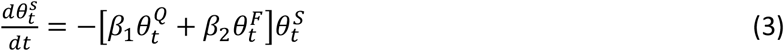

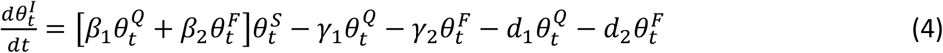

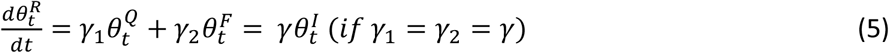

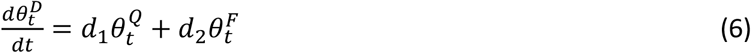

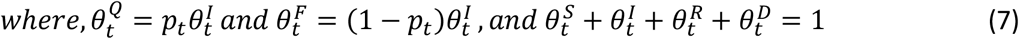

Here, 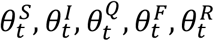 and 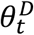 are the true but unobserved (latent) prevalence of susceptibles, infecteds, infected and quarantined, infected and undetected (free), recovered, and deceased respectively. In other words, they are the probabilities of a person being in the respective compartments at time *t*. Also, let 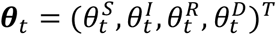 be the latent population prevalence.

Solution of this set of differential equations can be obtained using Runge-Kutta approximation. Let *f*(*θ*_*t*−1_,*β,γ*,***d***) denotes the solution of the set of differential equations for time *t*, where the function takes the values of the vectors *θ*_*t*−1_, ***β*** = (*β*_1_,*β*_2_)^*T*^, ***d***= (*d*_1_,*d*_2_)^*T*^and ***γ*** = (*γ*_1_,*γ*_2_)^*T*^ as the arguments. Then the fourth order Runge-Kutta approximation for the solution of these differential equations can be expressed as follows.

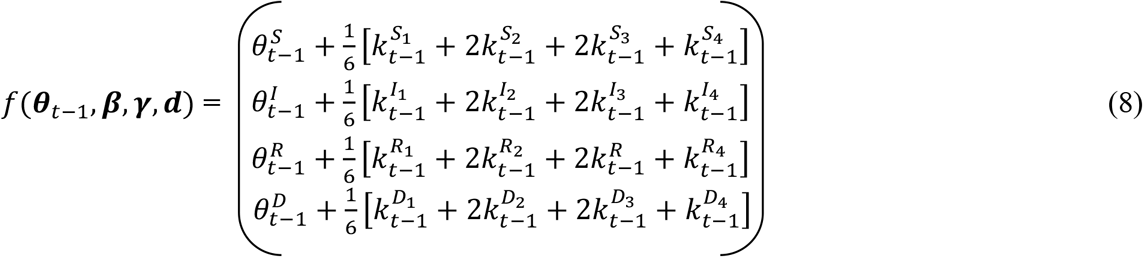

Complete expressions for 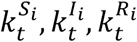 and 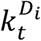 are presented in Appendix-A.

### 2.4. Dirichlet-Beta state-space formulation of the SI(Q/F)RD model

To account for the uncertainties in the epidemiological parameters and the transmission dynamics of the epidemic, we define a flexible state-space probabilistic model based on the deterministic SI(Q/F)RD model. Osthus *et al*. [13] introduced a Dirichlet-Beta state-space model based on the basic SIR model. We have extended the Dirichlet-Beta state-space model in accordance with the SI(Q/F)RD structure to estimate the model parameters, and forecast the transmission dynamics of the epidemic. Let 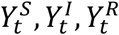 and 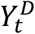 be the observed proportion of susceptibles, infecteds, recovered and deceased respectively. Then the Bayesian hierarchical state-space SI(Q/F)RD model can be defined as follows.

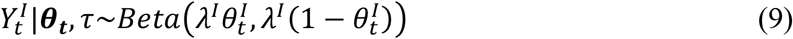

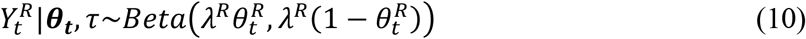

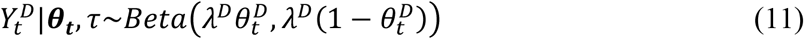

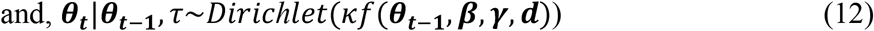

where, τ = {*θ*_**0**_,κ,*β,γ*,***d***,*λ*^*I*^,*λ*^*R*^,*λ*^*D*^}, *θ*_0_ is the baseline value of the vector ***θ***_***t***_, and *λ*^*I*^,*λ*^*R*^,*λ*^*D*^,κ > 0 control the variances of the distributions defined in equations (9), (10), (11), and (12) respectively. All other notations have already been defined in the section 2.3.

From equation (12) it is apparent that ***θ***_***t***_,*t*= 1,2 …, *T*, is a first-order markov chain. Also, the equations (9), (10) and (11) suggest that, for 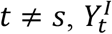 is independent of 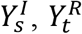 is independent of 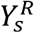, and 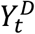 is independent of 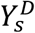, given 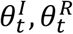 and 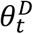 respectively.

Prior distributions of the model parameters can be defined as follows.

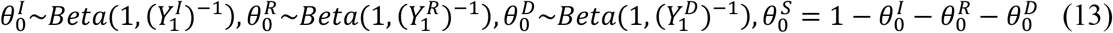

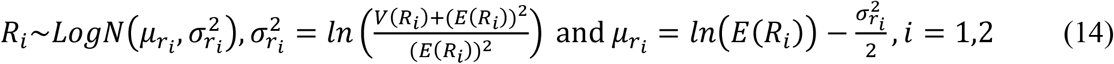

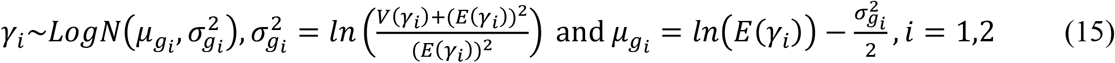

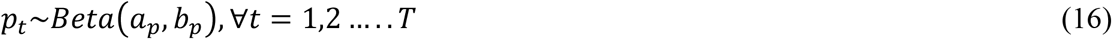

*R*_*1*_ and *R*_*2*_ are basic (average) reproduction rates associated with quarantined (Q) and undetected (F) infecteds, respectively. That is, 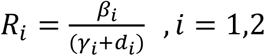.

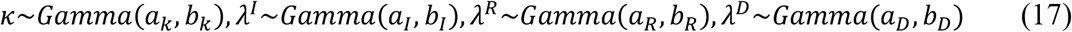

The hyperparameters of these Gamma prior distributions can be assumed according to the size of variability to be allowed in the Beta and Dirichlet distributions defined in equations (9)-(12). The higher the values of the parameters, κ,*λ*^*I*^,*λ*^*R*^ and *λ*^*D*^, the lower will be the variance of the respective Beta and Dirichlet distributions. If limited prior information is available regarding these parameters, a relatively flat Gamma prior distribution with a high expected value and a relatively higher variability is assumed while choosing the values of the hyperparameters *a’s* and *b’s*. Hyperparameters of the prior distribution of *p*_*t*_ are obtained from method of moments using the mean and variance of the daily estimates of underreporting. Hyperparameters of the lognormal distributions defined in the expressions (14) and (15) can be either based on historical knowledge on a similar epidemic, or can be estimated based on the observed data. In our study, we have used a time-series SIR (TSIR) model-based technique to estimate the hyperparameter for transmission rate, ***β*** [14]. Values for the hyper parameters ***γ*** and ***d*** are calculated using required information from published literature on COVID-19. *γ*_1_ = *γ*_2_ = *γ* (*say*) is taken as the inverse of the average recovery period. We have assumed equal recovery rates for both groups, quarantined and undetected. The components of ***d***, *viz*., *d*_*1*_ and *d*_*2*_ are estimated as, 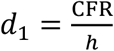 and 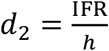, where *h* is the average number of days from infection till death and IFR is the infection fatality rate. CFR is the ratio of the number of deaths divided by the number of confirmed cases of disease. While, IFR is the ratio of deaths divided by the number of actual infections with SARS-CoV-2 and is generally expected to be lower than CFR.

### 2.5. Estimation of the hyperparameter *β* using TSIR model

The two components of the vector *β* = (*β*_1_,*β*_2_)^*T*^ must be estimated separately, such that they conform to their definitions. To do so, we have made some assumptions about the reported data, based on certain practical considerations. If it is known that the government of the country or the state under consideration did not take any significant action to quarantine infecteds or to promote physical distancing during the initial period of the epidemic, the transmission rate observed during that period can be safely considered as an initial estimate of *β*_2_ (the transmission rate due to infecteds who are not quarantined). Once the containment measures are imposed, the transmission rate based on the reported data is expected to change (reduce) and average transmission rate observed over the entire period of reporting can be considered as a safe initial estimate (hyperparameter estimate) for *β* (the overall average transmission rate as a result of both quarantined and undetected infecteds). This logic can be implemented using the TSIR model to estimate the hyperparameters as follows.

In TSIR model, the response, being a count variable, is assumed to follow certain discrete count process distribution like Poisson distribution or Negative Binomial distribution. The basic structure of TSIR model can be defined as follows; refer [15-17].

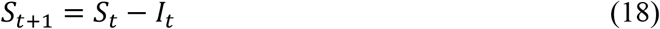

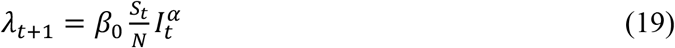

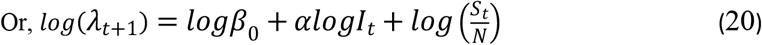

where, *S*_*t*_ and *I*_*t*_ are the number of susceptibles and infecteds (or infectives) at time *t, N* is the population size, *β*_*0*_ is the transmission rate and *λ*_*t*+1_ is the expected number of new infecteds at time *t+*1. New number of infecteds is assumed to follow Negative Binomial (or Poisson) distribution and a generalized Negative Binomial (or Poisson) linear model with log link is fitted with *logI*_*t*_ as a covariate and 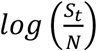as an offset variable. The exponent *α* is expected to be just under 1 (*i.e*., close to 1) and is meant to account for discretizing the underlying continuous process. However, we use an alternative interpretation of *α* based on the basic SIR model defined in equation (21). This method is drawn from our prior work where we have proposed a new method for obtaining time-varying estimates of transmission rate using TSIR model [14]. It is to be noted that the transmission rate is assumed to be time-varying, and hence, denoted as *β*_*t*_.

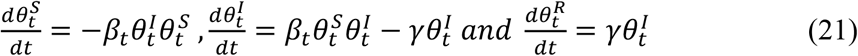

Using (21), the expression for expected number of new infecteds at time *t+*1 (taking *α* = 1) with a time-varying transmission rate can be written as follows.

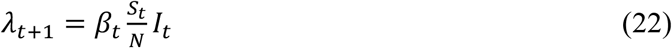

Comparing equations (19) and (22), we can see that if *α* = 1 (or close to 1), *β*_*t*_ *= β*_*0*_ (constant over time). However, if the value of *α* deviates considerably from 1, it has impact on the effective value of transmission rate, thus making the effective rate of transmission time-dependent. That is, in such cases *α* assimilates the empirical changes in transmission rate over time. Further, using equations (19) and (22), we can write,

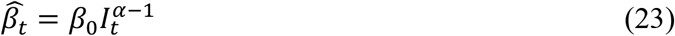

Now, suppose *T*_*1*_ represents the initial period of the epidemic, when proper quarantine protocols were not in place, and *T* represents the entire period for which the reported data on the epidemic is available. The estimates of *α* and *β*_*0*_ obtained from the fitting of the TSIR model shall be used in equation (23) to find estimates of transmission rate at each time *t*, 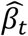, *t* = 1,2,3 …, *T*. Average of these estimates over a time period will give us the estimate of average transmission rate for that period. That is, the estimates of the transmission rates will be taken as, 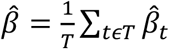 and 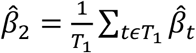. Then a reliable estimate of *β*_1_, the transmission rate due to the infecteds who are quarantined, can be obtained using the relation, 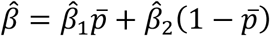; *where* 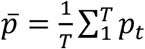. As a simpler, but logical, alternative to this step for finding 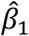, we can make use of the fact that the infecteds who are quarantined are expected to spread infection for, approximately, only one-third of the duration as compared to those who are not quarantined. This is because quarantined patients spread infections mostly in the incubation period of around 4-5 days, prior to getting quarantined, while infecteds who are not quarantined are expected to spread infection for the entire average infectious period of 14 days. So, after estimating 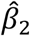 using the method described above, we can take 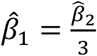, as the initial estimate.

### 2.6. Estimating parameters of the state-space SI(Q/F)RD model and forecasting

Posterior realizations on the parameters of the state-space model are generated using Gibbs sampling MCMC approach. The model is adopted in JAGS format and implemented in R using the package R2jags. Mean of posterior realizations of a parameter is taken as its posterior estimate. Further, 0.025 and 0.975 quantiles of the posterior realizations are taken as the limits of 95% credible intervals (CI) of the posterior estimates. Let *t*_*0*_ be the time till which the observations are available, and suppose that we wish to forecast the values of observed process 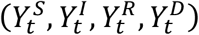 from *t*_*0*_+1 till the time *T*. We follow the following iterative steps to achieve our goal.

a. We generate *L* posterior realizations on the latent prevalence process *θ*_*t*_^(*l*)^,*l* = 1,2 …, *L* using Gibbs sampling approach, at each time point *t* = *t*_*0*_ +1,2…..,*T*. Here *L* is a sufficiently large number, say 1000 or more.
b. At each *t* (= *t*_*0*_ +1,2…..,*T*), and at each posterior realization of the prevalence process *θ* _*t*_^(*l*)^,*l* = 1,2 …, *L*, values of the observed process, say 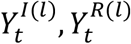 and 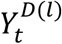 are simulated from their conditional distributions, 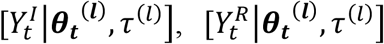 and 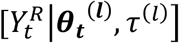,which are defined in the equations (9), (10) and (11), respectively.
c. Further, using the posterior realizations of *p*_*t*_^(*l*)^, at each *l* and each *t*, 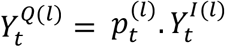 and 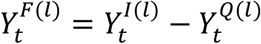 are also obtained. At each *t*, mean of the *L* simulated values serves as the estimate (forecasted value) of the respective variable (compartment proportion). 95% credible interval of each variable, at each time t, is also obtained using the 0.025 and 0.975 quantiles of the *L* values.

## 3. Implementation and Results

### 3.1. Data

Daily time-series data on total confirmed cases and total deaths for the states California and Florida are obtained from the github repository of the Centre for Systems Science and Engineering (CSSE), Johns Hopkins University, Maryland, USA (https://github.com/CSSEGISandData/COVID-19). Daily time-series data till 11 July 2020 was available at the time of procurement of data, and the same has been used for the entire analyses. Data on weekly state-wise estimates of excess deaths associated with COVID-19 till 11 July 2020, calculated as a difference between expected and reported number of deaths from all causes, is obtained from the website of CDC (https://www.cdc.gov/nchs/nvss/vsrr/covid19/excess_deaths.html). Data on rates of positivity of COVID-19 testing for the two states, California and Florida, are obtained from the official website of Johns Hopkins University on 29 July 2020 (https://coronavirus.jhu.edu/testing/testing-positivity).

### 3.2. CFR and data calibration to account for underreporting

To avoid the initial period of uncertain reporting due to the absence of sufficient government measures and proper reporting system at place, we have used the data from 02 March 2020 onwards for all analyses. The weekly excess death estimates are used to reconstruct the daily time-series data of deaths using the procedure discussed in section 2.1. According to the results reported by Verity *et al*. [18] based on patient level data from mainland China, the mean duration from onset of symptoms to death is 17.8 days (95% credible interval 16.9–19.2). They reported the best estimate of case fatality ratio in China as 1.38% (1.23-1.53), with substantially higher CFR in older age groups (0.32% [0.27–0.38] in those aged <60 years *vs* 6.4% [5.7–7.2] in those aged ≥60 years), up to 13.4% (11.2–15.9) in those aged 80 years or older). Their estimate for overall IFR in China is 0.66% [0.39-1.33], with an increasing profile with age. Yang *et al*. [19] reported that the median time from symptom onset to radiological confirmation of pneumonia is 5 days (interquartile range [IQR] 3–7 days); from symptom onset to intensive care unit (ICU) admission is 11 days (IQR 7–14 days); and from ICU admission to death is 7 days (IQR 3–11 days). That is, the estimate of median time from onset of symptoms to death can be taken as 11 + 7 = 18 days. This estimate of time-to-event of death is consistent with the results of Verity *et al*. [18]. So according to these results, the estimate of average duration from onset of infection to death should be around 17.8 + 5 ≈ 23 days, where 5 days is the average incubation period. However, for estimating delayed CFR based on the reported data and for estimating actual number of infecteds from the calibrated data on number of deaths, we have taken average duration from infected/ reported to death as 18 days. This is because infecteds are generally tested on the onset of symptoms, *i.e*., after the incubation period.

The delayed CFR values calculated based on reported data came out to be quite high in both states. For California, the delayed CFR was 3.36%, while for Florida it came out to be 3.17% (Table 1). Both estimates are quite higher than 1.38%, the estimate based on individual patient data as reported by Verity *et al*. [18]. In fact, some other reports have suggested even lower actual CFR of COVID-19. The Centre for Evidence-Based Medicine (CEBM) at the University of Oxford currently estimates the CFR globally at 0.51%, with all the caveats pertaining thereto; refer (https://www.virology.ws/2020/04/05/infection-fatality-rate-a-critical-missing-piece-for-managing-covid-19/). Since the estimate of CFR based on follow up data of individual patients is deemed as most reliable, especially during initial stages of the epidemic, as a conservative estimate, we have taken 1.38% as the standard CFR due to COVID-19. Thus, the results of CFR based on the reported data point at underreporting of number of infecteds in both states. Also, significantly high rates of positivity of COVID-19 testing in the two states-7.47% in California and 18.96% in Florida-as compared to the recommended rate of less than or equal to 5%, indicates lack of adequate amount of random testing. That is, it corroborates our assumption that the current testing strategy employed by the two states largely aim at targeted testing.

**Table 1:** **Delayed CFR estimates for the two states based on the reported data.**

| State | Total deaths till<br>16 July 2020 | Total confirmed till<br>29 June 2020 | Average Delayed CFR |
| --- | --- | --- | --- |
| California | 7535 | 223931 | 0.0336 (3.36%) |
| Florida | 4802 | 151389 | 0.0317 (3.17%) |

Using a delay of 18 days (between detection of infection and death), and a CFR of 1.38%, we employ the method discussed in section 2.2 to estimate the actual number of daily infected cases (new cases). It should be noted that due to the lag of 18 days in the formula, number of daily infected cases could be calculated till 24 June 2020 only (18 days prior to 11 July 2020). Data on number of recovered people is not reported by the two states. So, we have calibrated the data for number of recovered cases using the logic that if 1.38% is the average CFR, 98.62% will be the average case recovery rate. Following formula is used to estimate the daily count of recoveries.

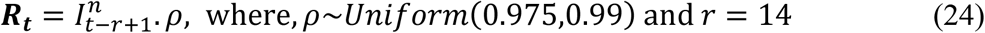

Here, 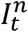 represents new number of infecteds at time *t*. The recovery rate, *ρ*, is randomly generated between 0.975 and 0.99 to introduce some amount of uncertainty into the calibrated data. The average duration of recovery, *r*, is taken as 14 days based on a WHO report [20]. However, at this juncture we should also note that some other studies have reported higher average duration of recovery from COVID-19. Verity *et al*. [18] have estimated mean duration from onset of symptoms to hospital discharge to be 24.7 days [95% CI: 22.9–28.1]. Recovery time also varies according to the severity of symptoms. Since small recovery time implies faster recovery rate, our choice of 14-day average recovery period (from the onset of symptoms) can be called as a conservative estimate, and the forecasts of transmission dynamics based on it can also be expected to be slightly on the conservative side. Since the formula given in equation (24) cannot give us the estimates of the first 13 days, we have constructed the daily recovery data for these initial days using daily recovery rate taken as the inverse of the average duration of recovery. Again, to induce some uncertainty in the data, we have randomly generated recovery rate, *φ*_*t*_, between 0.042 (1/24) and 0.071 (1/14), for each day. Following formula has been applied to implement the idea.

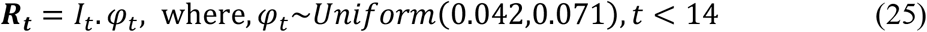

*I*_*t*_ is the number of (active) infecteds at time t. For *t < 14*, the already calculated values of recovered cases at time *t + 13* using equation (24), ***R***_***t+13***_, is adjusted by subtracting ***R***_***t***_ from it.

For further analyses, the reconstructed/ calibrated data on number of cases in each compartment is treated as the true data. The ratio of reported number of infecteds and true number of infecteds gives us the estimates of daily proportion of reporting *p*_*t*_ (*Q*_*t*_ *= p*_*t*_. *I*_*t*_). The calibrated data is provided in Appendix-B. Graphs of LOESS smoothed calibrated data on total number of deaths due to COVID-19 for the two states are presented in Figure 2. Summary statistics of *p*_*t*_ are provided in Table 2.

**Table 2:** **Summary statistics of *p*_*t*_, proportion of cases reported out of actual number of cases, obtained from the calibrated data.**

| Summary | California |  | Florida |  |
| --- | --- | --- | --- | --- |
| $p_t$ values (proportion of reporting) | Mean | Variance | Mean | Variance |
| First 30 days of the observed period | 0.034<br>(3.4%) | 0.0015 | 0.061<br>(6.1%) | 0.008 |
| Last 10 days of the observed period | 0.57<br>(57%) | 0.04 | 0.39<br>(39%) | 0.007 |
| Values lying between the first and the third sample quartiles | 0.26<br>(26%) | 0.01 | 0.23<br>(23%) | 0.007 |

**Figure 2:**
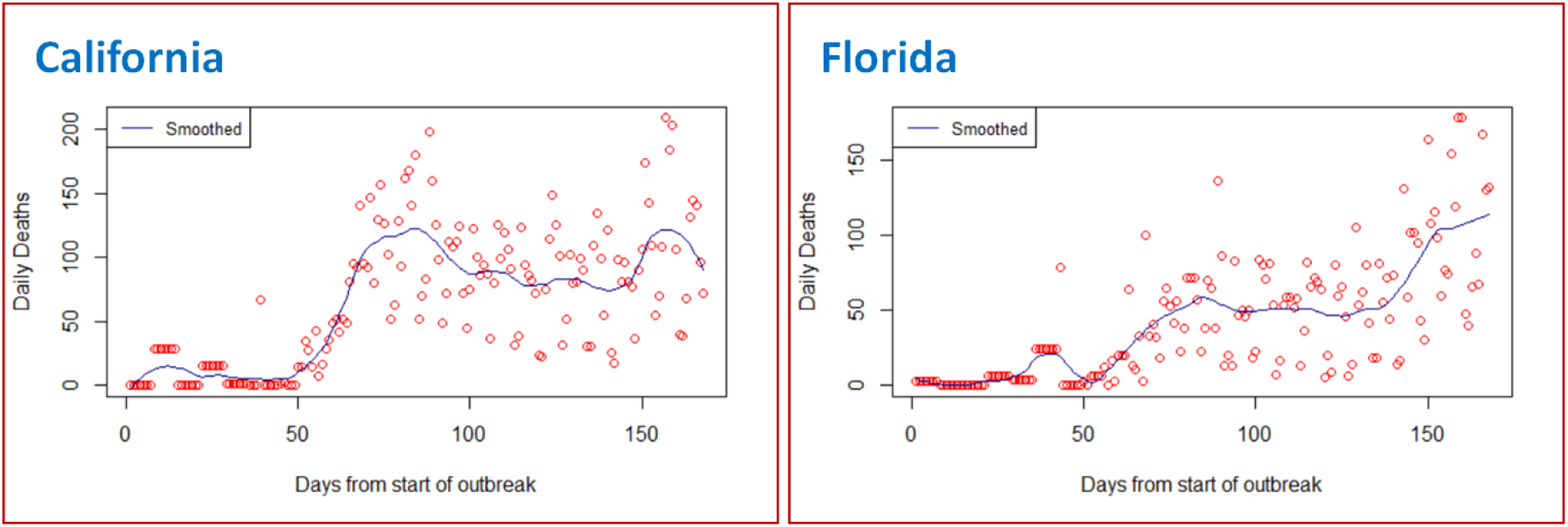
**LOESS smoothed calibrated data on total number of deaths due to COVID-19. Red circles represent the calibrated data points.**

### 3.3. Evaluating parameters and hyperparameters of the state-space SI(Q/F)RD model

In California, the first official lockdown measure was implemented on 19 March 2020, while in Florida it was implemented from 01 April 2020. So, the period till 18 March 2020 is considered as initial period of transmission for California, while the period till 31 March 2020 is taken as the initial period of transmission in Florida, for finding initial estimate of the transmission rate in the absence of proper quarantine measures for infecteds 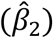. TSIR model described in section 2.5 is fitted assuming both Poisson and Negative Binomial distributions for the count process. Because of lower model deviance, model with Negative Binomial distribution is chosen over the Poisson model. These models are fitted in IBM-SPSS version 24. The estimated coefficients of the Negative Binomial TSIR models [model defined in equation (20)], for both states, are provided in Table 3. Using these coefficient estimates in equation (23) and taking average over respective initial periods of the COVID-19 epidemic, 02 March 2020-18 March 2020 for California and 02 March 2020-31 March 2020 for Florida, we have obtained the estimates of *β*_2_ for the two states. Initial estimate of *β*_1_ is calculated as one-third of the estimate of *β*_2_. These estimates are also provided in Table 3.

**Table 3:**
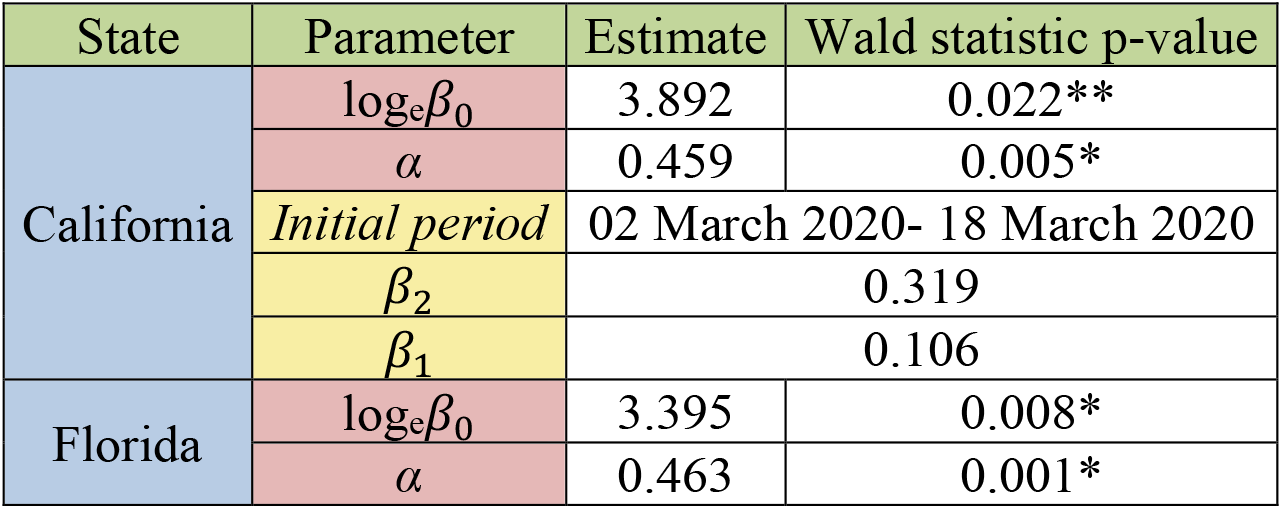
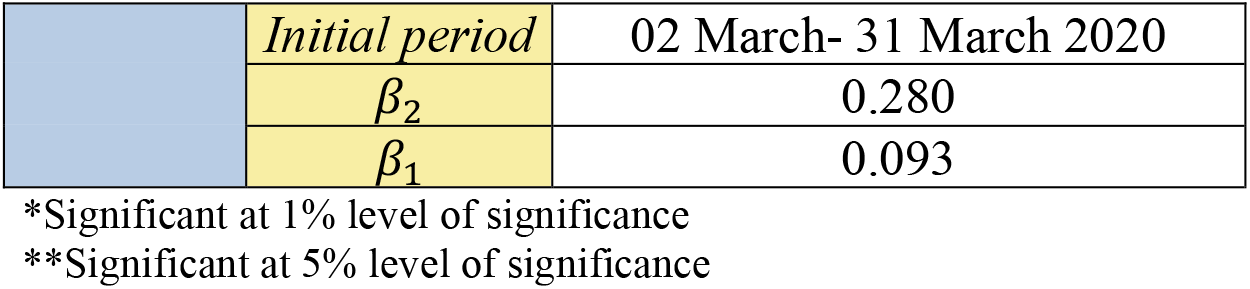
**Estimated coefficients of TSIR models with respective p-values of the Wald Chi-square statistic, and resultant estimates of *β*_*φ*_ and *β*_*φ*_. Based on the p-values of the Wald-test, all coefficient estimates are significant.**

| State | Parameter | Estimate | Wald statistic p-value |
| --- | --- | --- | --- |
| California | $\log_e \beta_0$ | 3.892 | 0.022** |
| | $\alpha$ | 0.459 | 0.005* |
|  | Initial period | 02 March 2020- 18 March 2020 |  |
| | $\beta_2$ | 0.319 | |
| | $\beta_1$ | 0.106 | |
| Florida | $\log_e \beta_0$ | 3.395 | 0.008* |
| | $\alpha$ | 0.463 | 0.001* |

**Table 3: Estimated coefficients of TSIR models with respective p-values of the Wald Chi-square statistic, and resultant estimates of $\beta_2$ and $\beta_1$ . Based on the p-values of the Wald-test, all coefficient estimates are significant.**
|  |  |  |
| --- | --- | --- |
|  | *Initial period* | 02 March- 31 March 2020 |
| | $\beta_2$ | 0.280 |
| | $\beta_1$ | 0.093 |

\*Significant at 1% level of significance
\*\*Significant at 5% level of significance

Using the estimates of CFR and IFR, and the average duration from onset of symptom to death, as reported by Verity *et al*. [18], we get the estimates of *d*_*1*_ and *d*_*2*_ as 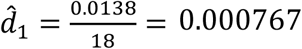 and 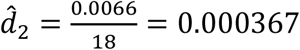.

Also, the estimate of recovery rate is 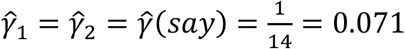. These estimates remain same for both states. So, the initial estimates of average reproduction numbers, *R*_*1*_ and *R*_*2*_, will be given as,

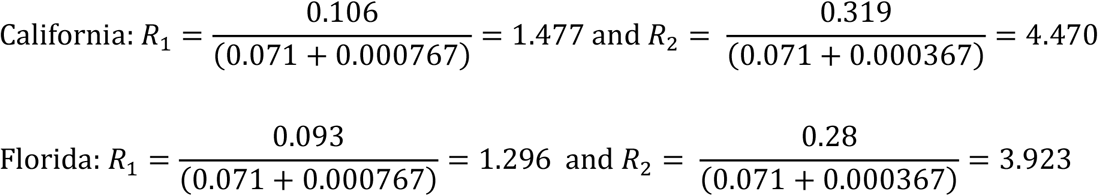

These estimates of *γ, R*_*1*_ and *R*_*2*_ are used to obtain informed hyperparameters of their prior distributions defined in equations (14) and (15). To decide on the hyperparameters of the Beta prior distribution of the time-varying proportion of quarantined infecteds, *p*_*t*,_ we use the descriptive statistics of its estimates over the observed period. Till the observed time period, the Beta prior distribution of *p*_*t*_ is assumed to have mean equal to 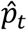, the estimate of *p*_*t*_ based on the calibrated data, and variance equal to the overall variance of the estimates. It is observed that after initial period of around one month, the values of 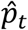 tend to first increase and then settle around some central value. So, for forecasting beyond the observed time period, sample mean and sample variance of the estimates corresponding to the last ten days of the observed period are taken as the mean and variance of the Beta prior distribution of *p*_*t*_. Complete list of prior distributions, along with the values of hyperparameters, used for fitting the state-space SI(Q/F)RD models for the two states is presented below.

California:

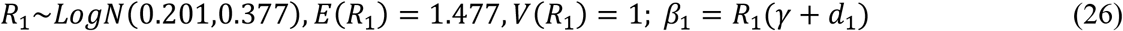

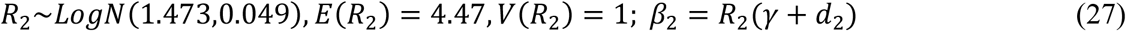

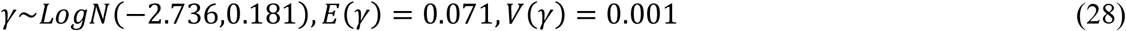

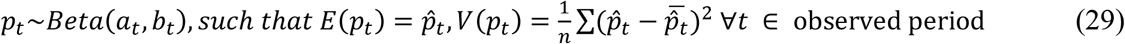

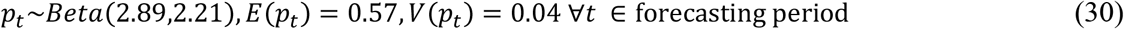

Florida:

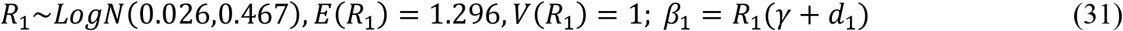

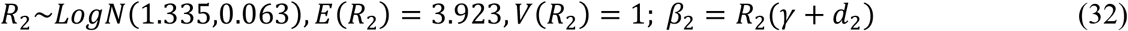

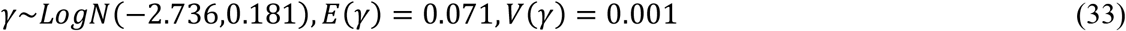

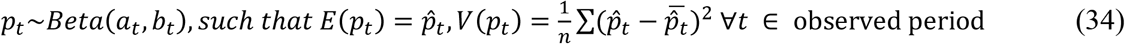

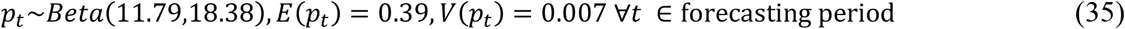

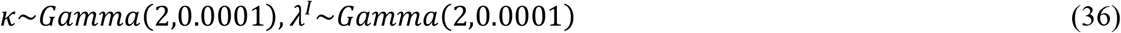

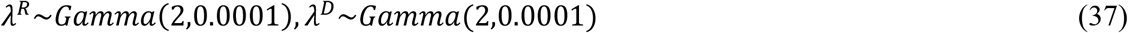

### 3.4. Posterior estimates and forecasts from the state-space SI(Q/F)RD model

The Dirichlet-Beta state-space SI(Q/F)RD model defined in section 2.4 is fitted on the calibrated data, using the parameters and hyperparameters obtained in section 3.3. The model is implemented in JAGS platform using R2jags package. Three parallel markov chains were run, each with 20,000 iterations of which first 10,000 were discarded. After thinning at an interval of 10, 1000 posterior simulations were saved from each chain, *i.e*., total 3000 posterior simulations were saved for each parameter. Posterior estimates of time-invariant parameters along with their standard deviations and 95% credible intervals for the two states are presented in Table 4 and Table 5. Plots of predicted values of the observed process on the number of infecteds 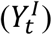 and the number of deaths 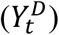, corresponding region of 95% credible intervals, and observed (calibrated) counts till the observed time, are exhibited for the two states in Figures 3 and 4. Total predicted size of the COVID-19 epidemic, in terms of number of infections and deaths, by the time it ends in the two states is presented in Table 6.

**Table 4:** **Posterior estimates of time-invariant parameters of the state-space SI(Q/F)RD model, along with their standard deviations and 95% credible intervals-California.**

| Parameter | Posterior mean | Posterior standard deviation | 95% credible interval |
| --- | --- | --- | --- |
| $R_1$ | 0.497 | 0.262 | [0.068, 1.004] |
| $R_2$ | 1.464 | 0.155 | [1.214, 1.813] |
| $\gamma$ | 0.069 | 0.006 | [0.056, 0.081] |
| $\kappa$ | 336063.593 | 47259.956 | [243264.879, 431918.329] |
| $\lambda^D$ | 1355.195 | 718.277 | [397.588, 2632.883] |
| $\lambda^I$ | 1012524.750 | 734717.729 | [1349.955, 2006982.462] |
| $\lambda^R$ | 1633152.503 | 334437.988 | [1073803.103, 2360304.964] |
| $\hat{\beta}_1 = \hat{R}_1(\hat{\gamma} + d_1)$ | 0.035 | | |
| $\hat{\beta}_2 = \hat{R}_2(\hat{\gamma} + d_2)$ | 0.102 | | |

**Table 5:** **Posterior estimates of time-invariant parameters of the state-space SI(Q/F)RD model, along with their standard deviations and 95% credible intervals-Florida.**

| Parameter | Posterior mean | Posterior standard deviation | 95% credible interval |
| --- | --- | --- | --- |
| $R_1$ | 0.359 | 0.224 | [0.052, 0.880] |
| $R_2$ | 1.612 | 0.097 | [1.416, 1.799] |
| $\gamma$ | 0.063 | 0.004 | [0.054, 0.071] |
| $\kappa$ | 500800.490 | 94547.445 | [327261.995, 679843.447] |
| $\lambda^D$ | 1022.341 | 303.916 | [539.044, 1629.331] |
| $\lambda^I$ | 999169.436 | 753473.727 | [4778.595, 2403835.884] |
| $\lambda^R$ | 1807366.511 | 365988.299 | [1164580.155, 2616920.665] |
| $\hat{\beta}_1 = \hat{R}_1(\hat{\gamma} + d_1)$ | 0.0229 | | |
| $\hat{\beta}_2 = \hat{R}_2(\hat{\gamma} + d_2)$ | 0.102 | | |

**Table 6:**
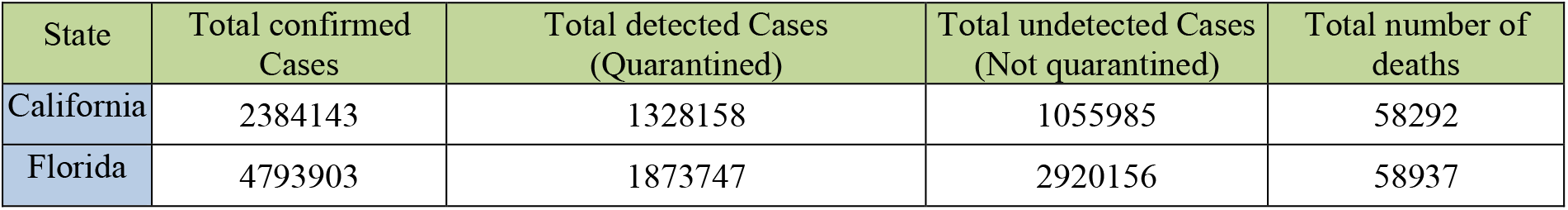
Predicted size of the COVID-19 epidemic in the two states.

**Figure 3:**
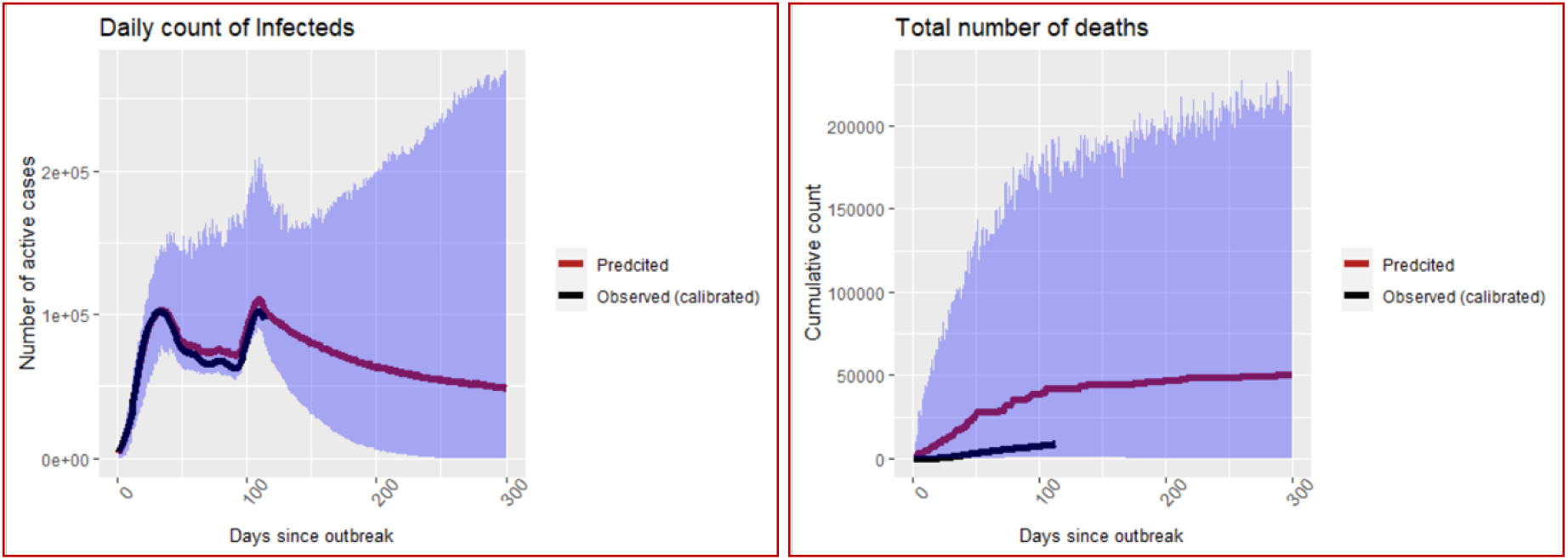
**Predictions of number of infecteds and number of deaths in California. The blue shaded ribbon is the region of 95% credible intervals.**

**Figure 4:**
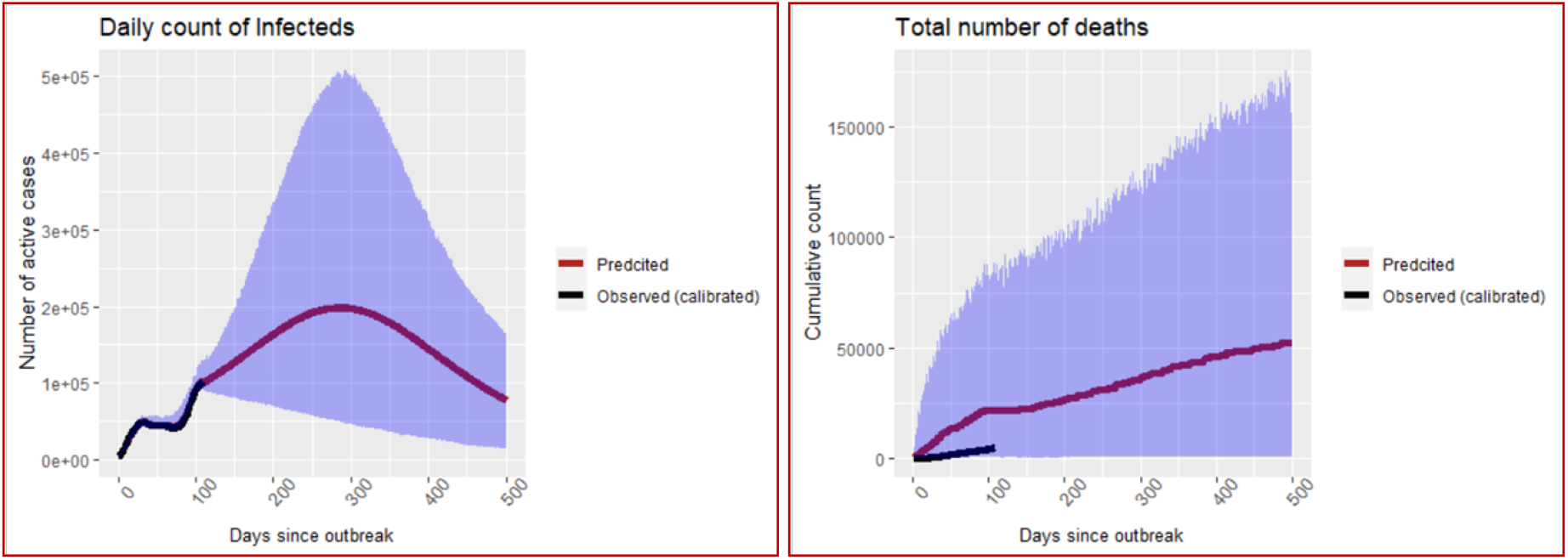
**Predictions of number of infecteds and number of deaths in Florida. The blue shaded ribbon is the region of 95% credible intervals.**

## 4. Discussion

### Some key insights from the results of the SI(Q/F)RD model

Estimates of the proportion of reported cases, *p*_*t*_, based on the calibrated data give us a good idea about the seriousness of the problem of underreporting. As can be seen from the summary provided in Table 2, the first 30 days of the outbreak of the COVID-19 epidemic have experienced extremely high level of underreporting (*i.e*., for the month of March as per the timeline of this study), with the average percentages of underreporting rate for the two states being 96.6% (California) and 93.9% (Florida). Similar results have been reported by Lau *et al*. [8]. Based on the reported data till 17 March 2020, they estimated the true number of infecteds to be 53.8 times higher than the reported number of cases in the United States, *i.e*., overall, 98.1% unreported cases. Similar trend was also reported by Wu *et al*. [7] and as per their findings the ratios of estimated cases to confirmed infections in California and Florida were around 17.5 (94.3% underreported cases) and 10 (90% unreported cases), respectively, till 18 April 2020. This can be attributed to the lack of appropriate testing facilities, including resources like testing kits, and absence of proper response from the government (or lack of seriousness and foresightedness on the part of the policy makers) during the initial stage of the pandemic. With time, the proportion of reported cases has increased steadily, and after a couple of months, it tends to fluctuate around the average values of 57% for California and 39% for Florida. These average values of *p*_*t*_, calculated at the later stage of the pandemic, are representative of the nature and capacity of testing policy of the states. For the same reason, the hyperparameters of the prior distributions of *p*_*t*_ for the purpose of forecasting are based on these later-stage averages. That is, due to the lack of extensive random testing in these states, around 43% infected cases in California and 61% infected cases in Florida, on an average, are expected to go unreported. These percentages of infecteds are undetected and are not quarantined, and they remain infectious for a much longer period than their quarantined counterparts, moving freely among the susceptibles. The SI(Q/F)RD epidemic model proposed by us is based on this hypothesis and the hypothesis is strongly supported by the posterior estimates of the transmission parameters obtained from the Dirichlet-Beta state-space SI(Q/F)RD model. Posterior estimates of average reproduction numbers associated with quarantined infecteds are 0.497 (sd: 0.262) and 0.359 (sd: 0.224), and for the undetected infecteds are 1.464 (sd: 0.155) and 1.612 (sd: 0.097) for California and Florida respectively. This clearly indicates that if almost all infecteds were quarantined; the number of active cases would have declined sharply, and the total size of the epidemic would have reduced drastically in both states. However, quarantining almost all infecteds, in the presence of a large proportion of asymptomatic cases, requires extensive amount of daily testing. This is clearly missing in both states under consideration, as indicated by very high rates of positivity of tests and high CFR values based on reported data for the two states.

### Comparing forecasted deaths with post-study published estimate of excess deaths

Since the true number of infecteds, detected plus undetected, remain latent in the population, it is not possible to compare forecasted values with the true values. However, comparing cumulative number of deaths forecasted by the state-space SI(Q/F)RD model with the estimated values of excess deaths can serve as a potent alternative to assess predictive efficiency of the model. Estimates of epidemiological parameters and predictions obtained from the state-space SI(Q/F)RD model in section 3 are based on the daily time-series data on number of cases reported till 11 July 2020 and the weekly estimates of excess deaths available till the same date. Predictive accuracy of the model will be determined by its ability to forecast true values beyond the training period of the model. From this perspective, we have plotted the forecasted time-series of cumulative number of deaths obtained from the fitted SI(Q/F)RD model along with the weekly estimated excess deaths due to COVID-19 till 14 November 2020. The estimates of excess deaths due to COVID-19 have been retrieved from the website of CDC (https://www.cdc.gov/nchs/nvss/vsrr/covid19/excess_deaths.html) on 4 December 2020. Striking difference in the estimated values of average reproduction numbers associated with detected (quarantined) cases and undetected cases suggest that the assumption regarding future values of proportion of detected cases plays a crucial role in ascertaining high predictive accuracy of the model. In other words, accuracy of the predictions from the proposed SI(Q/F)RD model relies greatly on the validity of the assumption regarding future testing policy in the region-*i.e*., on whether the testing capacity relative to the number of true cases is expected to increase, remain unchanged or decrease over time. To stress upon this argument, observed time-series of rates of positivity of COVID-19 tests are also plotted alongside the comparative plots of the forecasted number of deaths. Figure 5 and Figure 6 present these plots for California and Florida, respectively. In both figures, the left panel shows the comparative trends of cumulative number of deaths (predicted and excess) and the right panel presents the rate of positive tests over time. The trend line in blue in the right panel shows the average percentage of tests that were positive over the last seven days, *i.e*., a seven-day moving average of percentage of positive tests. The time-series plots of rates of positive tests for the two states are sourced from the website of Johns Hopkins University on 08 December 2020 [https://coronavirus.jhu.edu/testing/testing-positivity].

**Figure 5:**
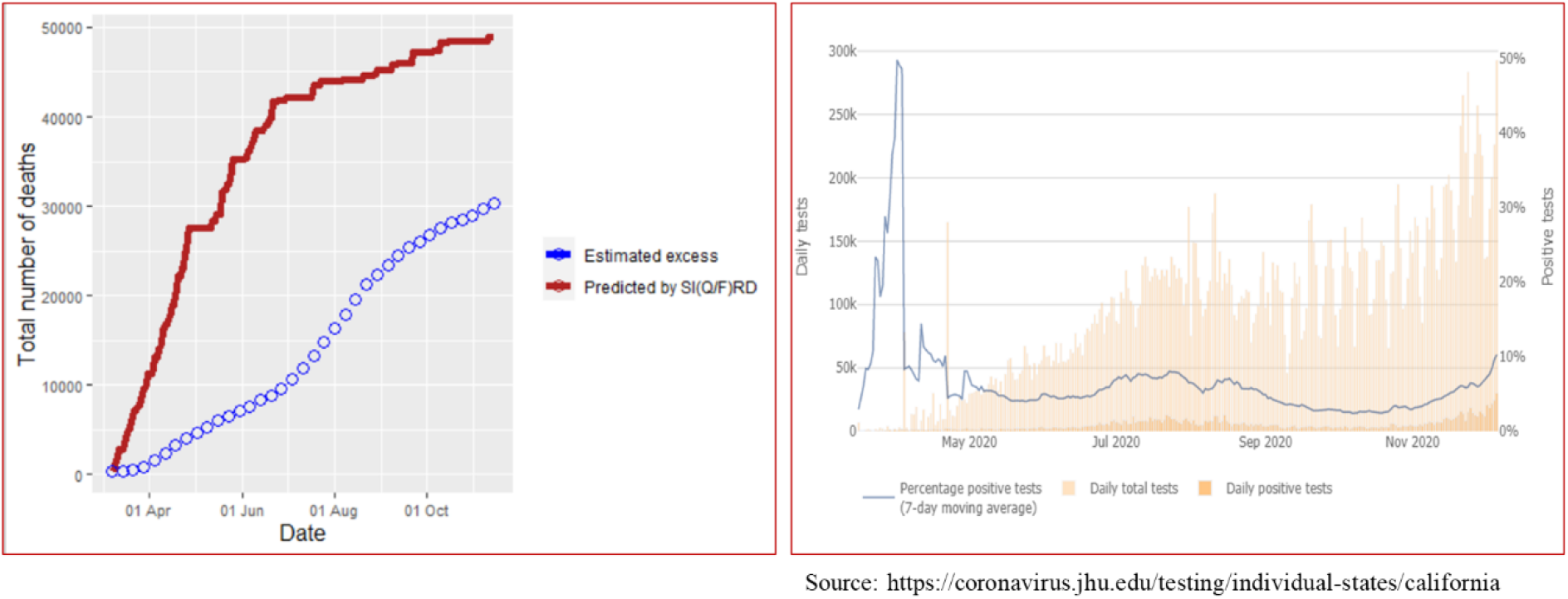
**California- Left panel shows comparison of cumulative number of deaths predicted by the SI(Q/F)RD model with the estimates excess deaths due to COVID-19. Right panel shows trend line of seven-day moving average of percentage of positive tests, along with daily total number of tests and daily total number of positive tests.**

**Figure 6:**
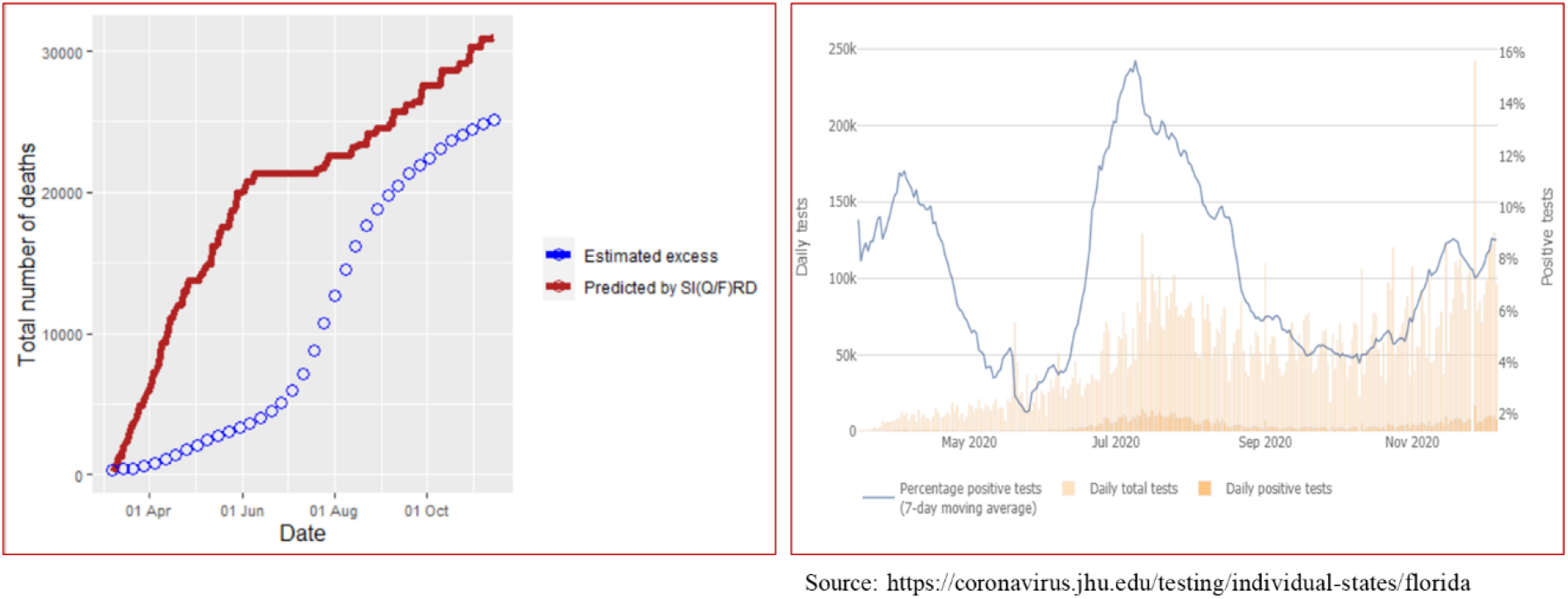
**Florida- Left panel shows comparison of cumulative number of deaths predicted by the SI(Q/F)RD model with the estimates excess deaths due to COVID-19. Right panel shows trend line of seven-day moving average of percentage of positive tests, along with daily total number of tests and daily total number of positive tests.**

In the case of California, there is a considerable difference between the predicted number of deaths and estimated excess deaths due to COVID-19. However, the difference is majorly in the scale of the values, and the two trend lines look similar in shape over time. Possible reason for the difference in the scale can be explained by analysing the trend line of rate of positive tests. The percentage of positive tests in California was extremely high during March-April, but although it started dropping exceptionally towards the end of April, it remained around 10% till July. However, September onwards the rate of positivity came down below 5%, the WHO recommended threshold. The steep rise in total number of tests performed daily, as shown by the pink towers in the graph, clearly explains this change. That is, California experienced a drastic change in testing capacity in the period of forecasting. Since, the hyperparameters of the model corresponding to the proportion of detected cases were defined based on the status of rate of positivity till July, the model tends to give overestimated forecasts of total number of deaths. Increasing the proportion of detected cases in the model as per the increase in testing capacity of the region would result in decrease in the total number of deaths. This is because the estimated rate of transmission for detected (quarantined) cases is relatively much lower than that of the undetected cases.

The scenario of rate of positive tests over time looks entirely different for Florida. Percentage of positive tests dipped below 5% for only two brief periods and it remained high for most of the time. That is, except for few short periods, the testing capacity has remained below par. Insufficient testing is also indicated by the fact that the trend line of the rate of positive tests is mostly parallel to the changes in the peaks of the total number of tests conducted per day. That is, it suggests that increase in the number of tests was not sufficient to reduce the rate of positive tests. Ideally, in the presence of sufficient amount of random testing, rate of positive tests should decrease with increase in the number of tests-as can be seen in the case of California. In other words, no significant change in the testing policy of Florida is observed in the forecasting period. This further implies that the hyperparameters defined for the proportion of detected cases in the state-space SI(Q/F)RD model remained valid for most of the forecasting period. Consequently, the forecasted values of cumulative number of deaths are much closer to the estimates of excess deaths in the case of Florida as compared to that of California. These results reaffirm the inevitable impact of testing capacity on the transmission dynamics of the pandemic, which forms the conceptual backbone of the proposed state-space SI(Q/F)RD model.

## 5. Conclusion

We have provided a comprehensive framework of data calibration and flexible epidemic modelling for forecasting the transmission dynamics of epidemics in the presence of serious underreporting of cases. The structure of the proposed SI(Q/F)RD model allows for adjusting the trajectory of the epidemic in terms of time-varying levels of underreporting. The Dirichlet-Beta state-space formulation of the SI(Q/F)RD model provides a dynamic approach to the estimation and prediction of both time-invariant and time-varying transmission parameters of the epidemic. Further, the proposed method, based on TSIR, for estimating hyperparameters of prior distributions of transmission rates (or reproduction rates) improves the posterior estimates by enriching the state-space model with strong prior information. Posterior estimates of the transmission parameters of the COVID-19 pandemic obtained for California and Florida emphasise on the need to incorporate different transmission rates for detected (quarantined) and undetected (not quarantined) cases in epidemic models. The findings also highlight the importance of extensive testing in the fight against pandemics like COVID-19. The proposed methodology will play an essential role in assessing and forecasting the true burden of any epidemic, irrespective of the level of underreporting of cases, even at a very early stage. Such reliable information is indispensable for policy makers for successful planning and implementation of containment measures.

### Limitations and further scope of research

We have taken the rate of death as a fixed (known) parameter in the state-space SI(Q/F)RD model. Finding its posterior estimates may improve the overall predictions from the model. Further, time varying transmission rates can be introduced in the model using the modifier functions to make the model more dynamic. The TSIR based method proposed for the construction of modifier functions by Deo *et al*. [14] can play an important role in implementing the SI(Q/F)RD state-space model with time-varying transmission rates.

## Data Availability

Data is procured from the github repository of Johns Hopkins University

https://github.com/CSSEGISandData/COVID-19

## Acknowledgements

We are extremely grateful to the reviewers and the editors for their invaluable comments and suggestions, which have helped us to greatly improve the paper.

## Appendix-A

### Expressions for 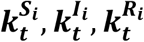 and 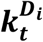

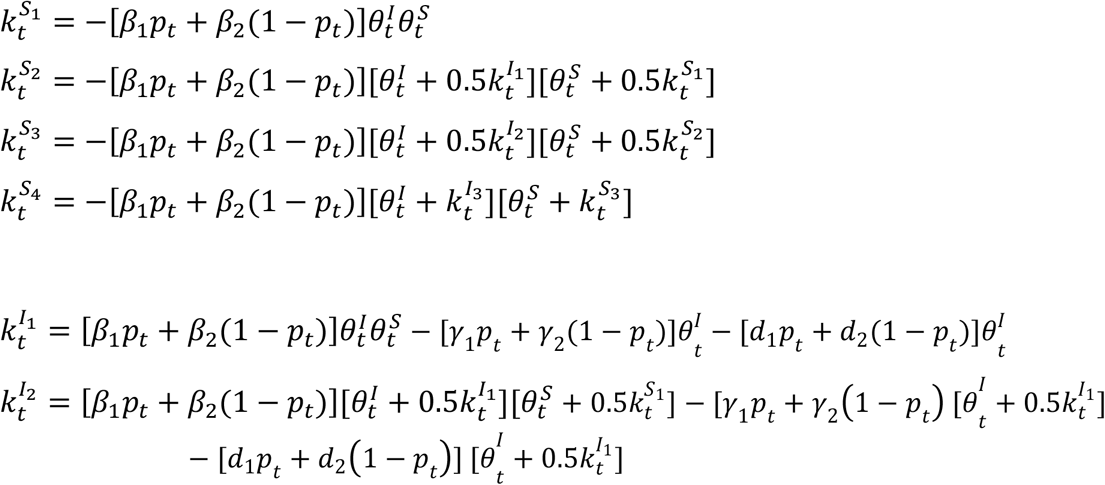

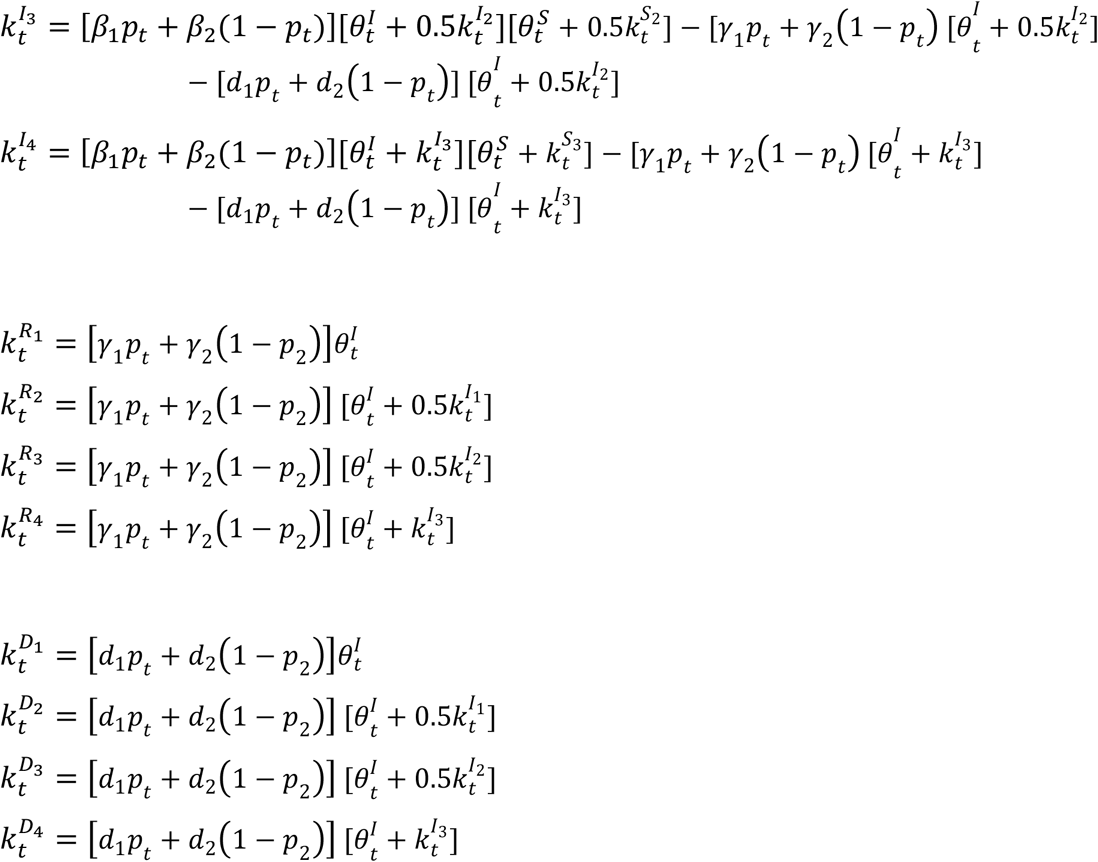

## Appendix-B

**Table B.1:** Calibrated data of California and Florida.

| Date | I_cali | p_obs_cali | R_cali | D_cali | I_florida | p_obs_florida | R_florida | D_florida |
| --- | --- | --- | --- | --- | --- | --- | --- | --- |
| 2-Mar-20 | 1449 | 0.0062 | 0 | 0 | 217 | 0.0046 | 0 | 0 |
| 3-Mar-20 | 2963 | 0.0025 | 80 | 0 | 569 | 0.0028 | 10 | 0 |
| 4-Mar-20 | 4575 | 0.0055 | 279 | 1 | 1115 | 0.0000 | 44 | 0 |
| 5-Mar-20 | 6337 | 0.0076 | 618 | 1 | 1785 | 0.0014 | 99 | 0 |
| 6-Mar-20 | 8300 | 0.0033 | 1046 | 1 | 2604 | 0.0000 | 222 | 0 |
| 7-Mar-20 | 10497 | 0.0080 | 1603 | 1 | 3565 | 0.0037 | 348 | 0 |
| 8-Mar-20 | 12817 | 0.0045 | 2399 | 1 | 4573 | 0.0024 | 570 | 2 |
| 9-Mar-20 | 15637 | 0.0017 | 3202 | 1 | 5727 | 0.0021 | 865 | 2 |
| 10-Mar-20 | 18592 | 0.0106 | 4304 | 2 | 6976 | 0.0012 | 1283 | 2 |
| 11-Mar-20 | 21862 | 0.0076 | 5525 | 4 | 8373 | 0.0050 | 1698 | 2 |
| 12-Mar-20 | 25183 | 0.0086 | 7204 | 4 | 9956 | 0.0030 | 2144 | 2 |
| 13-Mar-20 | 28787 | 0.0109 | 9107 | 4 | 11429 | 0.0092 | 2845 | 2 |
| 14-Mar-20 | 32367 | 0.0149 | 11606 | 12 | 13140 | 0.0134 | 3447 | 8 |
| 15-Mar-20 | 37606 | 0.0003 | 13024 | 22 | 15458 | 0.0142 | 3661 | 12 |
| 16-Mar-20 | 43209 | 0.0256 | 14583 | 34 | 17781 | 0.0004 | 4016 | 15 |
| 17-Mar-20 | 48952 | 0.0190 | 16361 | 49 | 20035 | 0.0315 | 4586 | 17 |
| 18-Mar-20 | 54635 | 0.0169 | 18415 | 66 | 22286 | 0.0390 | 5304 | 19 |
| 19-Mar-20 | 60181 | 0.0225 | 20748 | 86 | 24405 | 0.0414 | 6225 | 22 |
| 20-Mar-20 | 65496 | 0.0291 | 23454 | 108 | 26514 | 0.0414 | 7299 | 27 |
| 21-Mar-20 | 70606 | 0.0206 | 26507 | 133 | 28550 | 0.0610 | 8516 | 35 |
| 22-Mar-20 | 75324 | 0.0287 | 30093 | 162 | 30523 | 0.0711 | 9939 | 45 |
| 23-Mar-20 | 79730 | 0.0550 | 34060 | 195 | 32345 | 0.0644 | 11582 | 58 |
| 24-Mar-20 | 83739 | 0.0506 | 38491 | 233 | 34097 | 0.0521 | 13366 | 73 |
| 25-Mar-20 | 87259 | 0.0551 | 43406 | 276 | 35714 | 0.0745 | 15355 | 90 |
| 26-Mar-20 | 90316 | 0.1063 | 48777 | 326 | 37266 | 0.1826 | 17479 | 110 |
| 27-Mar-20 | 92797 | 0.0907 | 54718 | 382 | 38727 | 0.1441 | 19763 | 133 |
| 28-Mar-20 | 94737 | 0.0518 | 61267 | 444 | 40056 | 0.2247 | 22250 | 158 |
| 29-Mar-20 | 96359 | 0.0866 | 68272 | 513 | 41370 | 0.1212 | 24894 | 186 |
| 30-Mar-20 | 97665 | 0.1474 | 75658 | 589 | 42624 | 0.3024 | 27668 | 216 |
| 31-Mar-20 | 98899 | 0.1209 | 83253 | 673 | 43860 | 0.3019 | 30603 | 248 |
| 1-Apr-20 | 99983 | 0.1321 | 90990 | 765 | 45097 | 0.0508 | 33606 | 283 |
| 2-Apr-20 | 100788 | 0.1561 | 98927 | 864 | 46213 | 0.4807 | 36728 | 320 |
| 3-Apr-20 | 101418 | 0.1398 | 106961 | 968 | 47172 | 0.3019 | 39933 | 359 |
| 4-Apr-20 | 101761 | 0.0952 | 115134 | 1075 | 47931 | 0.3097 | 43263 | 400 |
| 5-Apr-20 | 101892 | 0.2591 | 123372 | 1184 | 48549 | 0.2006 | 46661 | 442 |
| 6-Apr-20 | 101726 | 0.1188 | 131760 | 1295 | 48994 | 0.2454 | 50158 | 486 |
| 7-Apr-20 | 101444 | 0.1673 | 140117 | 1408 | 49321 | 0.3138 | 53699 | 531 |
| 8-Apr-20 | 100961 | 0.1992 | 148456 | 1523 | 49510 | 0.2377 | 57304 | 578 |
| 9-Apr-20 | 100134 | 0.1276 | 156848 | 1639 | 49521 | 0.2418 | 61013 | 626 |
| 10-Apr-20 | 99040 | 0.1842 | 165289 | 1756 | 49375 | 0.3163 | 64806 | 675 |
| 11-Apr-20 | 97575 | 0.0892 | 173811 | 1873 | 49005 | 0.2664 | 68749 | 725 |
| 12-Apr-20 | 95891 | 0.1546 | 182407 | 1990 | 48586 | 0.3872 | 72740 | 776 |
| 13-Apr-20 | 93912 | 0.1659 | 191153 | 2107 | 47958 | 0.3171 | 76867 | 828 |
| 14-Apr-20 | 91738 | 0.2182 | 199876 | 2225 | 47240 | 0.1721 | 81083 | 881 |
| 15-Apr-20 | 89508 | 0.1989 | 208508 | 2345 | 46617 | 0.2445 | 85274 | 936 |
| 16-Apr-20 | 87084 | 0.1554 | 217188 | 2466 | 46076 | 0.2310 | 89382 | 992 |
| 17-Apr-20 | 84806 | 0.2348 | 225647 | 2589 | 45608 | 0.3925 | 93415 | 1050 |
| 18-Apr-20 | 82661 | 0.2148 | 233973 | 2712 | 45212 | 0.2034 | 97375 | 1109 |
| 19-Apr-20 | 80747 | 0.1443 | 242142 | 2834 | 44843 | 0.2277 | 101308 | 1168 |
| 20-Apr-20 | 78971 | 0.3497 | 250246 | 2955 | 44550 | 0.2065 | 105166 | 1226 |
| 21-Apr-20 | 77486 | 0.2782 | 258061 | 3074 | 44387 | 0.2200 | 108968 | 1283 |
| 22-Apr-20 | 76352 | 0.2870 | 265600 | 3191 | 44347 | 0.1201 | 112648 | 1339 |
| 23-Apr-20 | 75310 | 0.3425 | 272976 | 3306 | 44346 | 0.3623 | 116290 | 1394 |
| 24-Apr-20 | 74567 | 0.2782 | 280055 | 3419 | 44416 | 0.2394 | 119862 | 1448 |
| 25-Apr-20 | 73975 | 0.1571 | 286986 | 3529 | 44515 | 0.0828 | 123406 | 1501 |
| 26-Apr-20 | 73507 | 0.1868 | 293725 | 3635 | 44690 | 0.1878 | 126875 | 1553 |
| 27-Apr-20 | 73263 | 0.2216 | 300243 | 3738 | 44826 | 0.1642 | 130384 | 1604 |
| 28-Apr-20 | 73103 | 0.1891 | 306608 | 3837 | 44889 | 0.1924 | 133967 | 1654 |
| 29-Apr-20 | 72922 | 0.4194 | 312851 | 3934 | 44950 | 0.0944 | 137552 | 1704 |
| 30-Apr-20 | 72685 | 0.2300 | 319007 | 4029 | 45022 | 0.1364 | 141127 | 1753 |
| 1-May-20 | 72232 | 0.3230 | 325238 | 4121 | 45119 | 0.2827 | 144677 | 1802 |
| 2-May-20 | 71641 | 0.2307 | 331464 | 4211 | 45201 | 0.2010 | 148241 | 1852 |
| 3-May-20 | 70847 | 0.2753 | 337822 | 4299 | 45305 | 0.1688 | 151783 | 1902 |
| 4-May-20 | 70073 | 0.1736 | 344161 | 4386 | 45224 | 0.2294 | 155437 | 1952 |
| 5-May-20 | 69274 | 0.4551 | 350525 | 4473 | 45120 | 0.1560 | 159042 | 2002 |
| 6-May-20 | 68479 | 0.3822 | 356884 | 4561 | 44915 | 0.1668 | 162675 | 2052 |
| 7-May-20 | 67732 | 0.2676 | 363267 | 4650 | 44626 | 0.2516 | 166320 | 2102 |
| 8-May-20 | 67050 | 0.2886 | 369585 | 4739 | 44336 | 0.1186 | 169965 | 2153 |
| 9-May-20 | 66509 | 0.4782 | 375833 | 4829 | 44040 | 0.2452 | 173616 | 2204 |
| 10-May-20 | 66016 | 0.1751 | 382107 | 4918 | 43772 | 0.1891 | 177239 | 2255 |
| 11-May-20 | 65641 | 0.2910 | 388335 | 5007 | 43427 | 0.1308 | 180866 | 2306 |
| 12-May-20 | 65506 | 0.2745 | 394395 | 5096 | 43081 | 0.2973 | 184494 | 2357 |
| 13-May-20 | 65504 | 0.3023 | 400323 | 5184 | 42752 | 0.1608 | 188105 | 2408 |
| 14-May-20 | 65774 | 0.3406 | 406052 | 5272 | 42473 | 0.2631 | 191739 | 2459 |
| 15-May-20 | 66173 | 0.2993 | 411653 | 5359 | 42178 | 0.2948 | 195389 | 2510 |
| 16-May-20 | 66646 | 0.3338 | 417182 | 5444 | 41961 | 0.2159 | 199033 | 2561 |
| 17-May-20 | 67003 | 0.2396 | 422756 | 5527 | 41882 | 0.2411 | 202612 | 2612 |
| 18-May-20 | 67391 | 0.2248 | 428301 | 5608 | 41990 | 0.2575 | 206076 | 2663 |
| 19-May-20 | 67805 | 0.3735 | 433822 | 5687 | 42240 | 0.1572 | 209471 | 2714 |
| 20-May-20 | 68022 | 0.3793 | 439469 | 5765 | 42530 | 0.1640 | 212826 | 2765 |
| 21-May-20 | 68096 | 0.3516 | 445114 | 5843 | 42850 | 0.3474 | 216152 | 2815 |
| 22-May-20 | 68012 | 0.3937 | 450772 | 5921 | 43158 | 0.2321 | 219491 | 2864 |
| 23-May-20 | 67762 | 0.4091 | 456524 | 5999 | 43452 | 0.2056 | 222845 | 2912 |
| 24-May-20 | 67327 | 0.2733 | 462387 | 6078 | 43880 | 0.2187 | 226138 | 2959 |
| 25-May-20 | 66834 | 0.4360 | 468308 | 6157 | 44476 | 0.2464 | 229408 | 3006 |
| 26-May-20 | 66245 | 0.5560 | 474252 | 6237 | 45269 | 0.1438 | 232698 | 3053 |
| 27-May-20 | 65542 | 0.2945 | 480236 | 6318 | 46133 | 0.1085 | 236062 | 3100 |
| 28-May-20 | 64809 | 0.5114 | 486249 | 6400 | 47244 | 0.1638 | 239398 | 3146 |
| 29-May-20 | 64206 | 0.5198 | 492204 | 6483 | 48481 | 0.2756 | 242825 | 3192 |
| 30-May-20 | 63743 | 0.5914 | 498091 | 6566 | 49851 | 0.2050 | 246337 | 3238 |
| 31-May-20 | 63316 | 0.3685 | 504014 | 6650 | 51376 | 0.1600 | 249910 | 3285 |
| 1-Jun-20 | 63010 | 0.4859 | 509961 | 6734 | 53119 | 0.1382 | 253555 | 3332 |
| 2-Jun-20 | 62889 | 0.4282 | 515795 | 6818 | 55189 | 0.1224 | 257162 | 3380 |
| 3-Jun-20 | 63160 | 0.3590 | 521455 | 6901 | 57441 | 0.2363 | 260803 | 3429 |
| 4-Jun-20 | 63862 | 0.4473 | 526974 | 6984 | 59937 | 0.2448 | 264416 | 3479 |
| 5-Jun-20 | 65093 | 0.5269 | 532472 | 7067 | 62685 | 0.2163 | 268066 | 3530 |
| 6-Jun-20 | 66892 | 0.3857 | 537910 | 7149 | 65729 | 0.2004 | 271783 | 3581 |
| 7-Jun-20 | 69221 | 0.2621 | 543327 | 7229 | 68933 | 0.1798 | 275629 | 3632 |
| 8-Jun-20 | 72006 | 0.3858 | 548652 | 7307 | 72167 | 0.1452 | 279663 | 3683 |
| 9-Jun-20 | 75149 | 0.3379 | 553910 | 7384 | 75406 | 0.1593 | 283837 | 3734 |
| 10-Jun-20 | 78417 | 0.3518 | 559189 | 7460 | 78494 | 0.1943 | 288233 | 3786 |
| 11-Jun-20 | 81683 | 0.4282 | 564543 | 7536 | 81414 | 0.2358 | 292868 | 3840 |
| 12-Jun-20 | 84997 | 0.3672 | 569922 | 7611 | 84132 | 0.2626 | 297702 | 3897 |
| 13-Jun-20 | 88244 | 0.3817 | 575442 | 7685 | 86654 | 0.3523 | 302730 | 3956 |
| 14-Jun-20 | 91418 | 0.2618 | 581035 | 7759 | 88890 | 0.2784 | 308041 | 4018 |
| 15-Jun-20 | 94383 | 0.3852 | 586763 | 7834 | 90919 | 0.2422 | 313628 | 4083 |
| 16-Jun-20 | 97061 | 0.3720 | 592705 | 7910 | 92781 | 0.3714 | 319452 | 4151 |
| 17-Jun-20 | 99374 | 0.4487 | 598866 | 7987 | 94497 | 0.3459 | 325491 | 4222 |
| 18-Jun-20 | 101044 | 0.5157 | 605523 | 8066 | 95961 | 0.4193 | 331851 | 4297 |
| 19-Jun-20 | 101933 | 0.4554 | 612742 | 8146 | 97128 | 0.4933 | 338576 | 4376 |
| 20-Jun-20 | 102081 | 0.5496 | 620482 | 8229 | 98159 | 0.5175 | 345506 | 4458 |
| 21-Jun-20 | 101643 | 0.4445 | 628514 | 8316 | 98881 | 0.4490 | 352742 | 4543 |
| 22-Jun-20 | 100482 | 0.8345 | 636900 | 8410 | 99599 | 0.3751 | 360051 | 4632 |
| 23-Jun-20 | 98871 | 0.9648 | 645367 | 8511 | 100247 | 0.4159 | 367497 | 4726 |
| 24-Jun-20 | 96689 | 0.6972 | 653963 | 8619 | 100906 | 0.6824 | 375001 | 4824 |

## References

[1] WHO. (2020). Transmission of SARS-CoV-2: implications for infection prevention precautions. Retrieved from https://www.who.int/news-room/commentaries/detail/transmission-of-sars-cov-2-implications-for-infection-prevention-precautions

[2] Byambasuren, O., Cardona, M., Bell, K., Clark, J., McLaws, M.-L., & Glasziou, P. (2020). Estimating the Extent of True Asymptomatic COVID-19 and Its Potential for Community Transmission: Systematic Review and Meta-Analysis (pre-print). MedRxiv. doi:10.1101/2020.05.10.20097543

[3] CDC. (2020). COVID-19 Pandemic Planning Scenarios. Retrieved from https://www.cdc.gov/coronavirus/2019-ncov/hcp/planning-scenarios.html

[4] WHO. (2020). WHO Director-General’s opening remarks at the media briefing on COVID-19 - 16 March 2020. Retrieved from https://www.who.int/dg/speeches/detail/who-director-general-s-opening-remarks-at-the-media-briefing-on-covid-1916-march-2020

[5] Ministry of Health, N. (2020, August 21). COVID-19 - Testing rates for ethnicity and DHB. Retrieved from https://www.health.govt.nz/our-work/diseases-and-conditions/covid-19-novel-coronavirus/covid-19-current-situation/covid-19-current-cases/covid-19-testing-rates-ethnicity-and-dhb

[6] JHU. (2020). WHICH U.S. STATES MEET WHO RECOMMENDED TESTING CRITERIA? Retrieved August 27, 2020, from Johns Hopkins University: https://coronavirus.jhu.edu/testing/testing-positivity

[7] Wu, S.L., Mertens, A.N., Crider, Y.S., Nguyen, A., Pokpongkiat, N.N., Djajadi, S., et al. (2020). Substantial Underestimation of SARS-CoV-2 Infection in the United States. Nature Communications. https://doi.org/10.1038/s41467-020-18272-4

[8] Lau, H., Khosrawipour, T., Kocbach, P., Ichii, H., Bania, J., & Khosrawipour, V. (2020). Evaluating the Massive Underreporting and Undertesting of COVID-19 Cases in Multiple Global Epicenters. Pulmonology, 1502. https://doi.org/10.1016/j.pulmoe.2020.05.015

[9] CDC. (2020). Excess Deaths Associated with COVID-19. Retrieved July 23, 2020, from https://www.cdc.gov/nchs/nvss/vsrr/covid19/excess_deaths.htm

[10] Weinberger, D. M., Chen, J., Cohen, T., Crawford, F. W., Mostashari, F., Olson, D., & al., e. (2020, July 1). Estimation of Excess Deaths Associated With the COVID-19 Pandemic in the United States, March to May 2020. The Journal of the American Medical Association, E1–E9. doi:10.1001/jamainternmed.2020.3391

[11] Rivera, R., Rosenbaum, J. E., & Quispe, W. (2020). Excess Mortality in the United States During the First Peak of the COVID-19 Pandemic. MedRxiv (preprint). doi:https://doi.org/10.1101/2020.05.04.20090324

[12] Atkins, K. E., Wenzel, N. S., Ndeffo-Mbah, M., Altice, F. L., Townsend, J. P., & Galvani, A. P. (2015). Under-reporting and case fatality estimates for emerging epidemics. BMJ, 350:h1115. doi: doi: 10.1136/BMJ.h1115

[13] Osthus, D., Hickmann, K. S., Caragea, P. C., Higdon, D., & Del Valle, S. Y. (2017). Forecasting seasonal influenza with a state-space SIR model. Annals of Applied Statistics, 11(1), 202–224. doi:10.1214/16-AOAS1000

[14] Deo, V., Chetiya, A. R., Deka, B., & Grover, G. (2020). Forecasting Transmission Dynamics of COVID-19 in India Under Containment Measures-A Time-Dependent State-Space SIR Approach. Statistics and Applications, 18(1), 157–180.

[15] Bjørnstad, O., Finkenstadt, B., & Grenfell, B. (2002). Dynamics of measles epidemics: Estimating scaling of transmission rates using a time series sir model. Ecological Monographs, 72(2), 169–184.

[16] Finkenstadt, B., Bjørnstad, O. N., & Grenfell, B. (2002). A stochastic model for extinction and recurrence of epidemics: Estimation and inference for measles outbreaks. Biostatistics, 3(4), 493–510.

[17] Grenfell, B. T., Bjørnstad, O. N., & Finkenstadt, B. F. (2002). Dynamics of measles epidemics: Scaling noise, determinism, and predictability with the tsir model. Ecological Monographs, 72(2), 185–202.

[18] Verity, R., Okell, L., Dorigatti, I., Winskill, P., Whittaker, C., & al., e. (2020, March 30). Estimates of the severity of coronavirus disease 2019: a model-based analysis. The Lancet Infectious Diseases. doi:https://doi.org/10.1016/S1473-3099(20)30243-7

[19] Yang, X., Yu, Y., Xu, J., Shu, H., Xia, J., & al., e. (2020). Clinical course and outcomes of critically ill patients with SARS-CoV-2 pneumonia in Wuhan, China: a single-centered, retrospective, observational study. The Lancet Respiratory Medice, 8, 475-481. doi:https://doi.org/10.1016/S2213-2600(20)30079-5

[20] WHO. (2020). Report of the WHO-China joint mission on coronavirus disease 2019 (COVID-19). Retrieved from https://www.who.int/publications-detail/report-of-the-who-china-joint-mission-on-coronavirus-disease-2019-(covid-19)

